# Proteomics-based clustering outperforms clinical clustering in identifying heart failure patient groups with distinct outcomes

**DOI:** 10.1101/2025.08.19.25333956

**Authors:** Marion van Vugt, Ruicong She, Isabella Kardys, Teun B Petersen, Marie de Bakker, K Martijn Akkerhuis, Kadir Caliskan, Olivier C Manintveld, Alicia Uijl, Jan van Ramshorst, Dimitris Rizopoulos, Victor AWM Umans, Eric Boersma, David E Lanfear, Folkert W Asselbergs, Jessica van Setten, A Floriaan Schmidt

**Author notes:** Authors contributed equally (Marion van Vugt, Ruicong She, Isabella Kardys). Authors contributed equally (David E Lanfear, Folkert W Asselbergs, Jessica van Setten, A Floriaan Schmidt).

## Abstract

**Background.:** Clustering of heart failure (HF) patients typically relies on clinical characteristics, which may not reflect underlying pathophysiology relevant for personalised medicine. We aimed to identify plasma protein profiles of HF patients with reduced ejection fraction (HFrEF).

**Methods.:** Using latent class analysis, we derived clusters based on 1) clinical characteristics, and 2) proteomics (SomaScan) from 379 HFrEF patients. Survival analysis assessed associations with major cardiovascular (CV) events (a composite of HF hospitalization, CV death, heart transplantation, and left ventricular assist device implantation), HF hospitalization, CV death, and all-cause mortality. To aid clinical understanding, we identified differentially expressed proteins and explored their druggability.

**Results.:** Clustering on clinical characteristics identified three patient clusters that did not differ in disease progression. Proteomics-based clustering identified three clusters associated with disease progression. Cluster 1 included younger patients with fewer comorbidities, whereas cluster 3 consisted of older patients with more atrial fibrillation and renal failure. Cluster 2 had intermediate values for most characteristics; medication use was similar across clusters. Over three years of follow-up, cluster 1 had few events. Compared to this cluster, cluster 2 had increased risk of major CV events (HR 2.31, 95%CI 1.23; 4.36) and HF hospitalization (HR 2.30, 95%CI 1.10; 4.78), but not of death. Cluster 3 had high event rates with HRs of 5.84 (major CV events), 6.50 (HF hospitalization), 8.58 (CV death), and 5.07 (all-cause mortality). Results were externally validated in 511 HFrEF patients. Twelve proteins were differentially expressed, including druggable targets CD2, GDF-15, ABO, IGFBP-1, IGFBP-2, and RNase1.

**Conclusions.:** Proteomics-based HFrEF clustering identified three clusters associated with distinct disease outcomes, undetected using clinical characteristics.

## Introduction

Heart failure (HF) burdens society in terms of lost healthy life years, economic costs, and significantly reduces patients’ quality of life. HF is a syndrome characterized by ventricular dysfunction, subclassified by the left ventricular ejection fraction (LVEF) as HF with reduced ejection fraction (HFrEF) for LVEF ≤ 40%, HF with mildly reduced ejection fraction (HFmrEF) for LVEF 41-49%, and HF with preserved ejection fraction (HFpEF) for LVEF ≥ 50^1^. While this subclassification has clear clinical utility, the cutoff values for these categories are highly debated and it is widely recognized that patients within the same HF subclass meaningfully differ in pathophysiology and aetiology^2^. The heterogeneity in HF patients is a potential cause of frequent and late-stage failure of drug trials for HF, where a subset of patients may benefit from treatment, but the majority does not show sufficient benefit to warrant market authorization^3^.

Previous studies have attempted to improve subclassification by combining clinical patient characteristics with unsupervised multivariate clustering to identify patients with a more homogenous presentation and indirectly increase the homogeneity of pathophysiology^4^. Proteins are fundamental elements in biological processes, and clustering based on proteins could therefore lead to subclasses with a similar underlying disease mechanism. For example, increased levels of the circulating N-terminal pro-B-type natriuretic peptide (NT-proBNP) are indicative of myocardial stretch^5^, with NT-BNP levels being used for cardiac diagnosis, prognosis, and therapy. Given that proteins are the targets of most drug compounds, clustering on plasma proteins may result in HF clusters with a more homogeneous response to treatment. Previous proteomics-based clustering analyses have sourced a limited number of 92 cardiovascular (CV) prioritized proteins^6^. Our previous investigation in a subset of the Bio-SHiFT study identified three clusters^6^, whereas another study using the same 92 proteins identified six clusters^7^. Incorporating additional proteins that are not prioritized based on cardiovascular relevance may enhance clustering performance regarding disease association and may help identify novel proteins relevant for cardiovascular disease. In our previous work we identified four clusters using a wide range of serially measured plasma proteins^8^, whereas in the current study we aim to develop an approach that could be more easily applied in clinical settings without requiring repeated measures. Finally, while previous studies have replicated findings by applying the same clustering method to different datasets resulting in two or more models (i.e., a model for each dataset), they have not externally validated the same clustering model in independent data, hence limiting the clinical applicability of their results.

In the current study we therefore directly compared an HFrEF clustering model based on clinical characteristics to a model based on 4,210 plasma proteins combining latent class analysis (LCA) with principal component analyses (PCA). We included 379 HFrEF patients from the Bio-SHiFT study and externally validated the proteomic algorithm in 511 HFrEF patients from the Henry Ford HF PharmacoGenomic Registry (HFPGR). Subsequently, we explored differences in disease progression risk between HFrEF clusters and investigated druggability of the proteins driving cluster membership. Finally, to maximise applicability, we developed an open-access application programming interface allowing for the usage of our externally validated clustering model for other patients.

## Methods

### Study populations

Bio-SHiFT is a prospective cohort study of stable, chronic HF patients from Erasmus Medical Center Rotterdam, and Noordwest Ziekenhuisgroep, Alkmaar, the Netherlands^9^. Chronic HF patients aged 18 years or older were included during their regular outpatient visits. The study was approved by the Erasmus MC medical ethics committee, complied with the Declaration of Helsinki, and registered at ClinicalTrials.gov (NCT01851538). The derived proteomics-based clustering model was externally validated in HFPGR, a prospective observational registry from Detroit, MI, USA, which included HF patients aged 18 years or older and diagnosed with HF as defined by the Framingham Heart Study^10,11^. Only HFrEF patients of European descent with SOMAscan data were included for validation. This study was approved by the Henry Ford Hospital review board and complied with the Declaration of Helsinki. All patients provided written informed consent at enrolment. In the current study, we considered I) major CV event (a composite of HF hospitalization, CV death, heart transplantation, and left ventricular assist device implantation); II) HF hospitalization; III) CV death; and IV) all-cause mortality. Details on the study design and definitions of the outcomes are described in the **Supplemental Methods** and **Table S1**.

### Plasma protein measurements

Blood samples of the BioSHiFT cohort were collected at baseline, processed within two hours after collection, and stored at -80°C for a median of 5.3 years (Q1 4.1; Q3 6.8). Proteomic analyses of EDTA plasma samples were performed using the aptamer-based proteomic SOMAscan platform^12^ and the quality control of the proteomics has been described previously^8,13^. In total, 4,210 modified aptamers were measured, excluding aptamers with non-human, not validated targets, or with lower affinity than a competing aptamer for the same protein. In the current study, we used blood samples drawn at the time of inclusion and summarized the high-dimensional data using PCA. The protein measurements were normalized to a mean of 0 and a standard deviation of 1; original values are reported in **Table S2**. Details on the processing and quality control of proteomics data for the validation cohort are provided in the **Supplemental Methods** and the clinical event committee that adjudicated the outcomes was blinded to the proteomic results.

### Statistical analyses

We used LCA to derive a clustering model assigning HFrEF patients with similar clinical or proteomic profiles to clusters. In contrast to other clustering methods, LCA provides a straightforward function rule, which allows for assigning cluster membership to out-of-sample participants. For clinical clustering, variables with more than 5% missingness, related variables, and non-baseline variables arising during follow-up (such as drug prescriptions or devices) were excluded. Continuous clinical variables age, body mass index, and mean arterial pressure were categorized based on clinically relevant cut-offs, such as the World Health Organization classifications of normal weight, overweight, and obese for body mass index (**Table S3**), and for the plasma proteins, the principal components (PCs) were categorized using quartiles. Identification of the optimal number of clusters and categorization is described in the **Supplemental Methods** and **Figure S1**. A genetic algorithm was employed to identify the subset of clinical variables used in LCA. In short, the genetic algorithm performs an exhaustive search for the most relevant clustering variables, comparing multiple subsets of variables simultaneously and by slightly mutating these subsets over many iterations^14,15^. For the clustering on plasma protein levels, the first 20 PCs were used, and a sensitivity analysis was performed using the first 30 PCs (**Figure S1**). Additionally, we combined the clinical clustering variables with the first 20 PCs to evaluate the potential improvement in clustering performance from integrating these features (**Supplemental Methods**). Another sensitivity analysis using latent profile analysis is described in the **Supplemental Methods**.

We next determined the contrasts between HFrEF clusters based on clinical characteristics (age, aetiology, and history of coronary artery disease [CAD], arrhythmia, hypertension, and smoking) and clusters based on the first 20 PCs of the plasma proteins. First, we evaluated the difference in baseline patient characteristics, followed by tests for the difference in the cumulative risk of a major CV event, HF hospitalization, CV death, and all-cause mortality, employing the log-rank test. Hazard ratios (HR) and 95% confidence intervals (CI) were calculated using Cox proportional hazards models, truncating follow-up at three years, and we calculated the c-statistic to assess the discriminative ability of these clusters.

### Annotation of differentially expressed proteins

To facilitate model explainability, we identified the differentially expressed proteins between the three proteomics-based clusters using the *limma* package in R. We corrected for multiple testing using the Benjamini-Hochberg procedure to control the false discovery rate while maintaining a balance between sensitivity and specificity. Proteins were considered significantly differentially expressed if they had an Benjamini-Hochberg adjusted p-value < 0.05 and a minimum absolute log_2_-fold-change of 1.0. Each protein was assigned to the cluster with the highest expression. To gain biological insight, we next explored pathway enrichment using the R package *clusterProfiler*, leveraging the gene ontology resource and applying a Benjamini-Hochberg adjusted p-value threshold of 0.05 on proteins with a minimum absolute log_2_-fold change of 0.7. Next, we benchmarked the differences in disease progression per cluster against the association of these individual proteins using a Cox regression model and presenting the protein-specific HRs (per standard deviation increase in protein value) and 95% CIs using the differentially expressed proteins, age, and sex. To further benchmark the cluster associations with disease progression, we compared these to the associations of canonical cardiovascular related proteins alpha-actinin, BAG3, CRP, IL-6, TNF-alpha, cardiac troponin I, TnTc, tropomyosin, titin, and vinculin. Proteins were defined “druggable” if a drug targeting the protein is being tested in a clinical trial, and “drugged” if the compound has received marketing authorization^16^ ChEMBL^17^ and the British National Formulary databases were queried to obtain information on the indications and side effects of compounds targeting these proteins.

### External validation of the proteomics-based clustering model

To validate our proteomics-based clustering model, we applied it directly to external data from HFPGR. Unlike prior studies that replicate their findings by applying the same clustering method to different datasets resulting in two or more clustering models, we derived a single clustering model in BioSHiFT to assign cluster membership externally. This approach aligns with potential clinical application, where clinicians aim to assign individual patients to clusters with distinct progression rates.

## Results

Bio-SHiFT included 382 HFrEF patients of whom 379 had baseline SomaScan measurements available and were used in this study. Patients were included a median of 4.21 years [Q1 1.61; Q3 9.61] after their chronic HF diagnosis, they were predominantly male (n=275, 73%), of European ethnicity (n=348, 93%) with a median age of 64 years [Q1 56; Q3 72]. Reduced LVEF and proteomics were obtained at baseline. Furthermore, 104 patients (28%) had a New York heart association (NYHA) class III/IV, 165 (48%) had a history of coronary artery disease, and the median NT-proBNP level was 1,237 pg/mL [Q1 458; Q3 2,463] (**Table 1**, **Table S4**).

**Table 1.** Baseline table of the derivation (BioSHiFT) and validation (HFPGR) cohorts.

|  | BioSHiFT (n = 379) | Missing (%) | HFPGR (n = 511) | Missing (%) |
| --- | --- | --- | --- | --- |
| Demographics |  |  |  |  |
| Male sex, n (%) | 275 (72.6) | 0 | 357 (69.9) | 0 |
| Age (years) (median [IQR]) | 64.00 [55.50, 72.00] | 0 | 72.00 [63.00, 79.00] | 0 |
| Risk factors |  |  |  |  |
| BMI (kg/m<sup>2</sup>) (median [IQR]) | 26.39 [24.00, 30.05] | 1.8 | 29.76 [25.67, 34.20] | 0.2 |
| Systolic blood pressure (mmHg) (median [IQR]) | 115.00 [100.00, 130.00] | 3.7 | 125.00 [111.50, 140.00] | 0 |
| Diastolic blood pressure (mmHg) (median [IQR]) | 70.00 [60.00, 78.00] | 3.7 | 69.00 [62.00, 78.00] | 0 |
| Mean arterial pressure (mmHg) (median [IQR]) | 85.33 [75.00, 93.33] | 3.7 | 88.33 [80.00, 97.83] | 0 |
| Hypertension, n (%) | 164 (43.7) | 1.1 | 434 (84.9) | 0 |
| Smoking, n (%) | 269 (71.4) | 0.5 | 344 (67.3) | 0 |
| HF-related measures |  |  |  |  |
| Etiology, n (%) |  |  |  |  |
| Exposure toxic substances | 20 (5.3) |  | 5 (1.0) | 0 |
| Hypertension | 33 (8.7) |  | 97 (19.0) | 0 |
| Ischemic heart disease | 163 (43.0) |  | 289 (56.6) | 0 |
| Unknown | 23 (6.1) |  | 90 (17.6) | 0 |
| Valvular disease | 12 (3.2) |  | 8 (1.6) | 0 |
| NYHA class = III/IV, n (%) | 104 (27.6) | 0.5 | 111 (78.3) | 0 |
| LVEF (%) (median [IQR]) | 30.00 [22.25, 37.00] | 21.4 | 36.00 [30.00, 45.00] | 0.8 |

| Clinical history |  |  |  |  |
| --- | --- | --- | --- | --- |
| CAD, n (%) | 165 (48.2) | 9.8 | 344 (67.3) | 0 |
| AF, n (%) | 135 (36.1) | 1.3 | 184 (36.0) | 0 |
| Arrhythmia, n (%) | 150 (39.9) | 0.8 | 42 (8.2) | 0 |
| Stroke, n (%) | 48 (12.8) | 1.3 | 41 (8.0) | 0 |
| Renal failure, n (%) | 179 (47.4) | 0.3 | 6 (1.2) | 0 |
| Diabetes, n (%) | 97 (25.6) | 0 | 194 (38.0) | 0 |
| Biomarkers |  |  |  |  |
| Creatinine (mg/dL) (median [IQR]) | 1.22 [1.01, 1.51] | 10.8 | 1.06 [0.83, 1.37] | 6.3 |
| NTproBNP (pg/mL) (median [IQR]) | 1,236.75 [458.41, 2,463.30] | 0.8 | 1,929.50 [886.12, 3,891.73] | 0 |
| eGFR (mL/min/1.73m<sup>2</sup>) (median [IQR]) | 56.97 [43.40, 73.79] | 10.8 | 65.33 [46.18, 86.72] | 6.3 |

Abbreviations: AF = atrial fibrillation, BMI = body mass index, CAD = coronary artery disease, eGFR = estimated glomerular filtration rate, HF = heart failure, HFPGR = Henry Ford HF Pharmacogenomic Registry, IQR = interquartile range, LVEF = left ventricular ejection fraction, NT- proBNP = N-terminal-pro hormone B-type natriuretic peptide, NYHA = New York Heart Association.

### Deriving HFrEF patient clustering models

Leveraging clinical information on the 379 BioSHiFT patients resulted in a model assigning patients to three clusters with an ischemic aetiology, hypertensive aetiology, or underlying cardiomyopathy (**Supplemental results**). Using the plasma proteomics resulted in a clustering model which similarly assigned patients to three clusters (**Figure S2CD**). The differences in patient characteristics were less evident compared to that of the clinical clusters. Proteomic cluster 1 consisted of the youngest patients with a median age of 59 years, lower NYHA class (83% I/II), and a lower comorbidity burden. Patients in proteomic cluster 3 were older with a median age of 71 years, 61% had a history of coronary artery disease, 50% of atrial fibrillation, and 70% suffered from renal failure (**Table S5**, **Figure S3**). No differences were observed in medication use among the proteomic clusters. The patient overlap between the clinical and proteomic clusters was limited (**Figure 1**). Deriving a “combined” clustering model combining clinical characteristic with plasma proteins resulted in a hybrid solution where the patients in cluster 1 of the proteomics and the combined model were nearly identical, while clusters 2 and 3 were mixed (**Table S6, Figure 1**).

**Figure 1.**
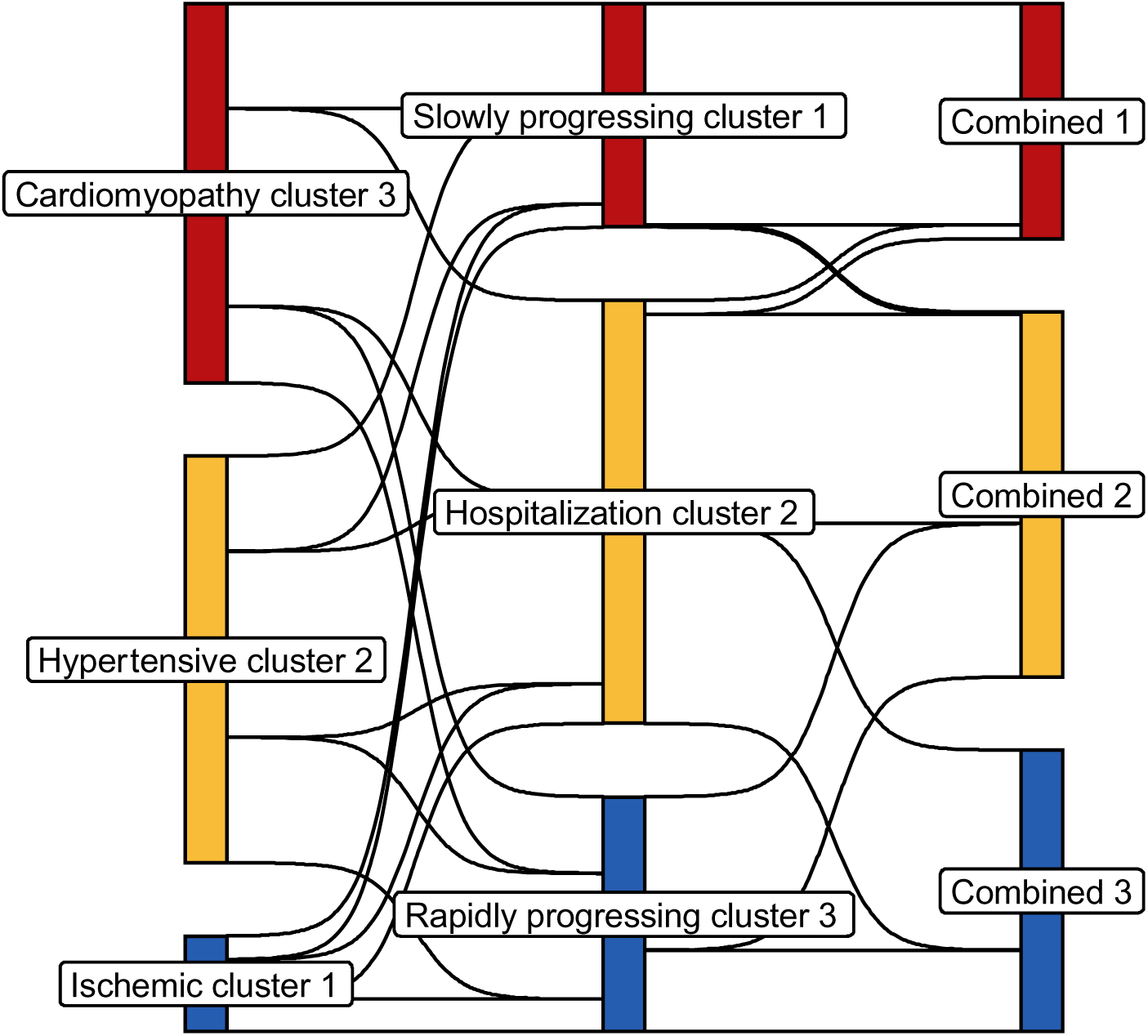
HFrEF patient flow between the different clustering models. N.B. The flow represents the proportion of HFrEF patients (n=379) that were assigned to different clusters in the various approaches. For example, this figure shows that the clinical clusters (i.e. ischemic cluster 1, hypertensive cluster 2, and cardiomyopathy cluster 3) are very different from the proteomic clusters, whereas the proteomic slowly progressing cluster 1 and combined cluster 1 are almost identical.

### Associating cluster membership with outcomes

During a median follow-up of 2.24 [Q1 1.39; Q3 2.60] years, 113 patients experienced a major CV event, 89 patients were hospitalized for HF, 41 patients died, of which 32 suffered a CV death (**Table S4**). Clinical cluster membership was not significantly associated with any of the clinical outcomes (**Supplemental results**, **Figure S4A, Table S7**). Protein cluster membership was significantly associated with major CV events, HF hospitalization, CV death, and all-cause mortality (**Figure 2**, p-value < 0.001 for all outcomes). The disease associations of the combined clustering model (i.e. using both the clinical characteristics and protein values) were attenuated relative to the model exclusively using the available plasma proteins (**Supplemental results**, **Figure S4B**, **Table S7**).

**Figure 2.**
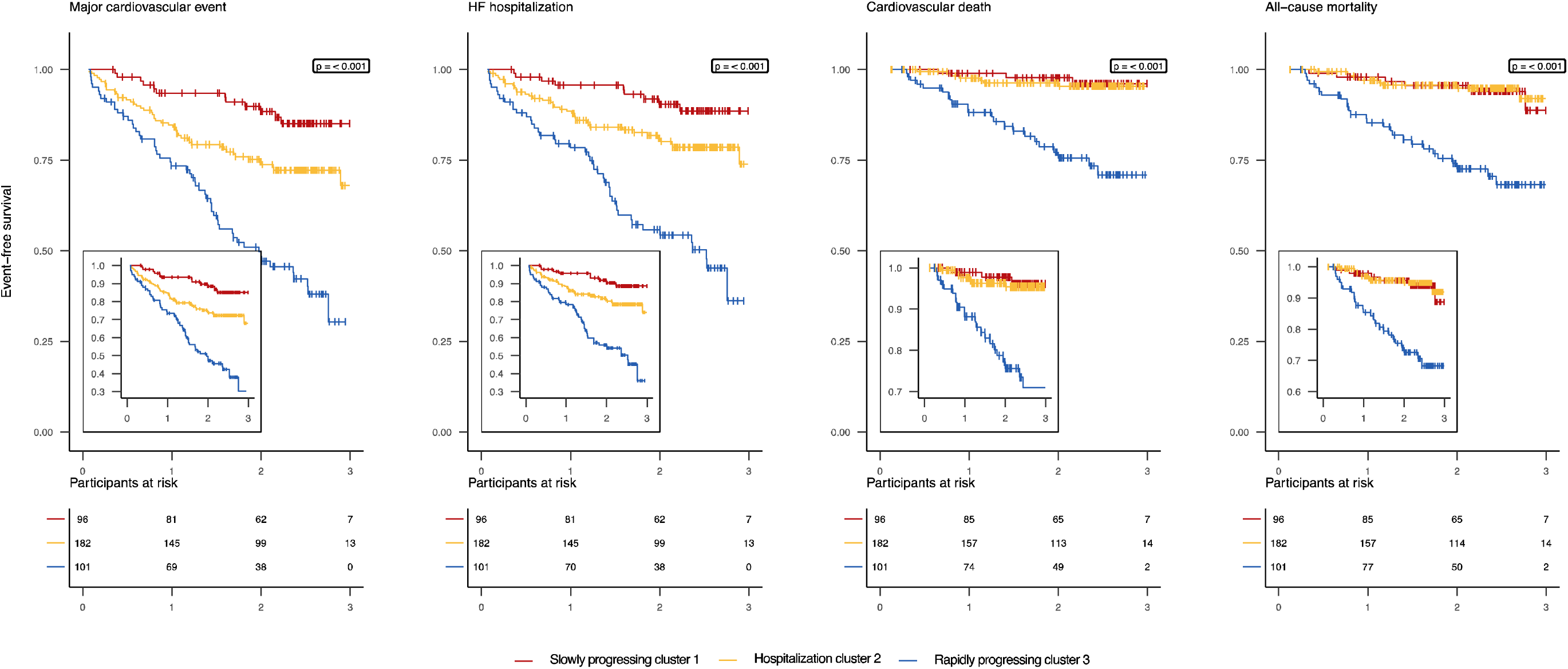
Event free survival is significantly different across proteomics-based clusters. N.B. p-values were calculated using the log-rank test. Abbreviations: HF = heart failure.

Focussing on the disease associations of the proteomics-based clustering model, we found that compared to the slowly progressing cluster 1, patients in proteomics-based cluster 2 had an increased rate of major CV events (HR 2.31 95% CI 1.23; 4.36) and HF hospitalizations (HR 2.30 95%CI 1.10; 4.78), which did not translate into an increased rate of fatal events (**Figure 3**). Patients in the rapidly progressing proteomics-based cluster 3 had a more pronounced increased event rate compared to patients in slowly progressing cluster 1: HR 5.84 (95%CI 3.12; 10.93) for major CV events, and HR 6.50 (95%CI 3.17; 13.33) for HF hospitalization, which did result in an increased rate of fatal events: HR 8.58 (95%CI 2.56; 28.67) for CV death and HR 5.07 (95%CI 2.08; 12.33) for all-cause mortality (**Figure 3**). The c-statistic for proteomics-based clustering ranged between 0.69-0.74 (**Table S7)**.

**Figure 3.**
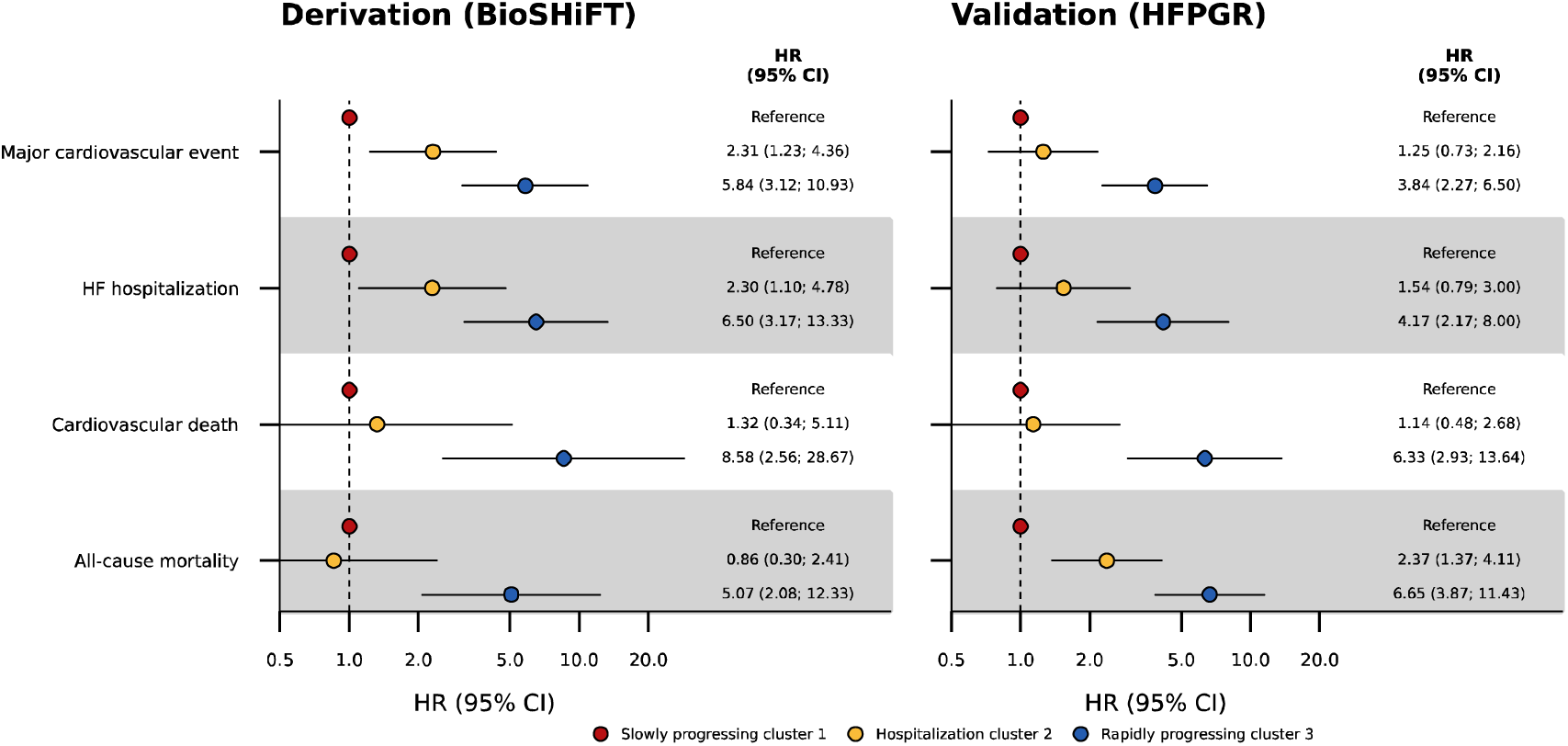
The association between proteomics-based cluster membership and disease progression in people with HFrEF. NB. Follow-up was truncated at three years. The model was derived in 379 participants of the BioSHiFT cohort and externally validated in 511 participants of the HFPGR. Bars represent the 95% confidence interval. Abbreviations: CI = confidence interval, HF = heart failure, HFPGR = Henry Ford HF PharmacoGenomic Registry, HR = hazard ratio.

A sensitivity analysis based on the first 30 protein PCs similarly resulted in three clusters with comparable outcome associations and latent profile analysis did not identify clusters with a similarly strong association with disease progression.

### Externally validating the proteomics-based clustering model

We next sought to externally validate the proteomics-based clustering model by replicating the associations between cluster membership and disease progression in 511 HFrEF patients from HFPGR. Patients were included a median of 6.03 years (Q1 1.98; Q3 10.87) after diagnosis with a median age of 72 years (Q1 63; Q3 79) and 69.9% males (**Table 1**, **Table S8**). Here we found that, relative to the slowly progressing cluster 1, patients in cluster 2 had an increased rate of all-cause mortality (HR 2.37, 95%CI 1.37; 4.11). Similar to the derivation cohort, patients in rapidly progressing cluster 3 were at an increased risk of major CV events (HR 3.84, 95%CI 2.27; 6.50), HF hospitalization (HR 4.17, 95%CI 2.17; 8.00), CV death (HR 6.33, 95%CI 2.93; 13.64), and all-cause mortality (HR 6.65, 95%CI 3.87; 11.43; **Figure 3, Table S9**).

### Interpreting clustering model and identifying potentially druggable proteins

Next, to improve model interpretability, we identified differentially expressed proteins across the three proteomics-based derived clusters. We observed that ABO and tumor protein p53-inducible protein 11 (TP53I11) values were significantly upregulated in the slowly progressing cluster 1, whereas the values of ten proteins were significantly upregulated in the rapidly progressing cluster 3 (showing intermediate high values in hospitalization cluster 2): peroxidasin homolog (PXDN), neuroblastoma suppressor of tumorigenicity 1 (NBL1), RNase1, insulin-like growth factor binding protein (IGFBP) 2, REG-3-alpha, growth-differentiation factor-15 (GDF-15), Apolipoprotein F, NT-proBNP, IGFBP-1, and CD2 (**Table S10)**. Filtering for a log_2_-fold-change of 0.7 resulted in 56 additional proteins and pathway enrichment analysis showed that slowly progressing cluster 1 was characterised by proteins involved in lipid metabolism (such as ADH4, APOA5, APOC3, and CRYZL1). Rapidly progressing cluster 3 was characterised by proteins involved in the immune response (such as EPHA2, RNase1, MMP12, NBL1, and REG-3-alpha) and extracellular matrix organisation (such as COL28A1, CST3, PRSS2, PXDN, and TNFRSF1A). Hospitalization cluster 2 was characterised by energy metabolism and glucose usage (**Table S11**).

To benchmark cluster-outcome associations, we compared the HRs (**Figure 3**) to the disease HRs of the differentially expressed protein values, supplemented by the HRs of ten canonical proteins (**Figure S5**). While NT-proBNP, IGFBP-2, GDF-15, NBL1, and PXDN were significantly associated with clinical outcomes, their individual associations were markedly smaller than the HRs for cluster membership (**Figure 3, Figure S5**).

Of the twelve differentially expressed proteins, CD2 is targeted by drug compounds alefacept and siplizumab, which are indicated for diabetes type I, plaque psoriasis, and arthritis. Additionally, GDF-15, ABO, IGFBP-1, IGFBP-2, and RNase1 are druggable. GDF-15 is inhibited by visugromab and ponsegromab which are evaluated in clinical phase oncological trials (**Table S12**).

## Discussion

In the current study, we found that a plasma protein-based model derived three HFrEF clusters with distinct outcome associations. In contrast, a clustering model based on clinical variables resulted in aetiological clusters which did not associate with HFrEF outcomes.

Furthermore, after externally validating our proteomics-based clustering model, we have made this publicly available for potential usage in clinical settings: https://gitlab.com/mvvugt/clusimp.

Patients in proteomics-based rapidly progressing cluster 3 had worse outcomes for all clinical outcomes compared to slowly progressing cluster 1 and hospitalization cluster 2, which was also observed in the external validation cohort. Interestingly, patients in hospitalization cluster 2 had worse outcomes for major CV events and HF hospitalization compared to those in slowly progressing cluster 1 but showed a comparable low rate of fatal events compared to slowly progressing cluster 1. Despite a strong association with disease progression, medication use was equal in all proteomics-based clusters, implying that patients are treated sub optimally and indicating the potential for personalized treatment based on their proteomics profile.

Out of the twelve differentially expressed proteins, eight have been previously associated with cardiac traits or diseases. Specifically, common genetic variants in the *ABO* gene have been associated with heart failure^18–21^ and myocardial infarction^22–24^, while variants in *TP53I11* have been associated with resting heart rate^25,26^ and hypertrophic cardiomyopathy^27^. These proteins were significantly upregulated in slowly progressing cluster 1, which was characterised by an altered lipid homeostasis. Hospitalization cluster 2 was intermediate in many aspects, such as expression of the proteins and characterised by glucose usage in the pathway analysis.

PXDN has previously been shown to be upregulated in HFrEF patients compared to healthy individuals^28^, where it plays a role in fibrosis and extracellular matrix formation. RNase1 has been proposed as HF biomarker due to its increased expression in HF patients and cardioprotective effects by catalysing RNA degradation, thereby mitigating inflammatory responses^29^. RNase1, NBL1, and REG-3-alpha are involved in immune response mechanisms and together with PXDN, these proteins and processes characterise the rapidly progressing cluster 3. Both IGFBP-1 and IGFBP-2 are involved in inflammation and metabolism, and modulate the effects of IGF-1, which is crucial for cellular development and survival. They are hypothesised to play a role in HF pathophysiology and have been suggested as biomarkers for mortality and CV disease risk^30–33^. The combination of high levels of IGFBP-1 and NT-proBNP have previously been associated with worse prognosis in HFrEF patients^7^. GDF-15 is a stress response protein highly expressed in cardiomyocytes upon several HF-related pathophysiological conditions such as inflammation, oxidative stress, hypoxia, and tissue injury. Compared to NT-proBNP, another stress biomarker in HF, GDF-15 is expressed in more tissues and therefore provides different information compared to NT-proBNP in a systemic disease such as HFrEF^34^. Both proteins have been associated with HF severity and prognosis^35–40^, with GDF-15 proposed as an HF drug target. Drugs are currently being developed for GDF-15, offering potential opportunities for repurposing, while other proteins such as ABO, IGFBP-1, IGFBP-2, and RNase1 present promising targets due to their druggability.

Benchmarking the association between cluster membership with disease progression against the individual associations of the differentially expressed proteins showed a substantially stronger association than for any individual protein (HR between 5.07 and 8.59 for rapidly progressing cluster 3, compared to an HR < 2 for any of the individual proteins). This included higher values of NT-proBNP and GDF-15, which showed an HR per standard deviation of at most 2. This suggests that the joint consideration of multiple protein values by the clustering model is essential to more accurately identify people with a meaningfully worse disease outcome.

### Limitations

We anticipate that significant survival differences for clinical clustering and the attenuated association for combined clustering may become evident in larger sample size settings. For example, a clustering study on 6,909 HFpEF patients applying similar methods, identified five clusters based on clinical characteristics which were associated with disease outcomes^41^. Hence, our results do not show and absence of differences between patients clustered using clinical characteristics or clinical characteristics combined with proteins but rather suggest that the difference is larger when using proteomics exclusively. Despite the improvement compared to clinical clustering, the c-statistics and high dimensional nature of the proteomic clustering limits immediate clinical applicability. Our findings represent a first step toward identifying biologically distinct HFrEF subgroups. Future work should aim to develop a minimal, robust set of proteins that can more accurately assign patients to clusters. These steps were not pursued in the present study to avoid overfitting due to repeated analyses on the same data. Future studies should also explore the contribution of the overexpressed proteins to cardiac metabolism and pathogenesis, adding to the known associations with cardiac disease. These proteins might also be useful in other clinical settings, for which clinically useful cut-off values should be established to guide individual patient management. However, in our study, cut-off values are not required because the clustering method assigns patients to clusters based on their overall proteomic profile. Our findings are based on proteomic data from one timepoint, a median of 4.21 years after initial diagnosis. Protein values are anticipated to be influenced by disease stage and concomitant medication usage at the time of proteomics measurement has potential implications for the derived clustering model. Some variation may reflect this biological and clinical heterogeneity, which is also representative of real-world clinical settings. Medication usage was broadly similar across the clusters, and thus unlikely to have caused the observed differences in associations with clinical outcomes. Importantly, we have externally validated the clustering model in a US healthcare setting where patients were recruited a median of 6.03 years after initial diagnosis. The storage duration of the blood samples may have affected the stability of the proteins. We expect the influence on our results to be limited, because storage duration was relatively consistent across patients. While protein degradation might have affected the detectability of individuals proteins and the set of differentially expressed proteins, the clustering approach is likely robust to this variation, because it is based on relative patterns across the proteome. Nevertheless, application of the clustering model in newly diagnoses HF patients deserves further careful consideration. This study, as well as previous clustering studies, has predominantly included male HF patients from European descent. While this reflects the higher risk of HFrEF observed in males^42–44^, this imbalance requires careful consideration regarding generalizability. To enhance the clinical applicability of our work, we limited the study to HFrEF patients and to support similar studies in HFpEF, HFmrEF, or any HF population, we have released the computational code. Due to the moderate sample size, we categorised continuous variables to simplify the model and reduce sample size requirements, which might have affected model expressivity. In larger sample size settings, more flexible algorithms may further improve the already highly discriminative model’s current performance. Nevertheless, the derived model was successfully validated in an external cohort that differed considerably from the derivation cohort, underscoring the robustness of our approach.

## Conclusions

We derived and externally validated a clustering model leveraging information on plasma proteins to identify three groups of HFrEF patients associated with distinct disease progression. These differences in disease progression were more pronounced when patients were clustered on clinical characteristic only. Cluster membership was driven by twelve differentially expressed proteins, of which six were drugged or druggable, providing potential leads for HF drug development.

## Acknowledgements

None

## Data availability statement

The clustering algorithm is available online at https://gitlab.com/mvvugt/clusimp. Anonymized data that support the findings of this study will be made available to other researchers for purposes of reproducing the results upon reasonable request and in accordance with a data-sharing agreement.

## Non-standard abbreviations and acronyms

CI: confidence interval
CV: cardiovascular
EF: ejection fraction
HF: heart failure
HFmrEF: HF with mildly reduced ejection fraction
HFpEF: HF with preserved ejection fraction
HFrEF: HF with reduced ejection fraction
HR: hazard ratio
IQR: interquartile range
LCA: latent class analysis
LV: left ventricular
NT-proBNP: N-terminal-pro hormone B-type natriuretic peptide
NYHA: New York heart association
PC: principal component
PCA: principal component analysis

