## Supplemental Material for "Proteomics-based clustering outperforms clinical clustering in identifying heart failure patient groups with distinct outcomes"

**Supplement**

**Supplemental Methods**

*Study designs and outcome definitions*

At baseline, information of Bio-SHiFT participants was collected on HF symptoms and aetiology, CV risk factors, medical history, and medication use, and electrocardiography and echocardiography were performed. Baseline and three-monthly follow-up visits included short medical examinations and the blood collection. All patients provided written informed consent. A clinical event committee determined the outcomes based on hospital records and discharge letters, without knowledge of the proteomic measurements.

In Bio-SHiFT, HF hospitalization was defined as hospitalized for an exacerbation of HF symptoms, together with two of the following conditions: a) brain natriuretic peptide or NT-proBNP more than three times the normal upper limit; b) signs of worsening HF, including pulmonary rales, increased jugular venous pressure or peripheral oedema; c) increased dose or intravenous administration of diuretics; or d) administration of positive inotropic agents. Death was considered due to a CV cause if the cause of death included myocardial infarction, ischemic heart disease, heart disease, HF, sudden cardiac death, sudden undefined death, unwitnessed death, ill-described death, or stroke.

In HFPGR, HF hospitalization was based on ICD9 and ICD10-codes, heart transplant and left ventricular assist device implantation on CPT codes, and mortality was sourced from the CDC – National Death Index (**Appendix Table S1**).

*Clustering variables and number of clusters*

Categorization of the principal components was done using different percentile-based cut-offs: quartiles (25th, 50th, 75th), deciles (10% intervals), as well as custom groupings based on the 10th–90th and 20th–80th percentiles to define low, middle, and high values.

Clustering was performed using all types of categorization and the optimal categorization was chosen based on the highest entropy.

Clusters were derived using maximum-likelihood estimation, with the optimal number of clusters identified using the Bootstrap Likelihood Ratio Test[1]. In short, we compared the *k* cluster model with the lowest Bayesian Information Criteria with a *k+1* cluster model calculating the likelihood ratio for the two models. The distribution of the likelihood ratio was estimated using 999 bootstraps, selecting the model with *k+1* clusters based on a p-value of 0.05 or smaller. Patients were assigned to clusters based on the highest probability of cluster membership.

As a sensitivity analysis we performed the proteomic clustering using latent profile analysis (LPA) using the R package *mclust*, which does not require categorization of the continuous principal components.

*Combined clustering*

In addition to clinical and proteomic clustering, we performed a combined clustering analysis by integrating clinical clustering variables with the first 20 principal components (PCs) derived from the proteomic variables. This approach aimed to determine if the combination of clinical and proteomic data improved patient stratification. The methodology was identical to that used for the clinical and proteomic clustering. We assessed differences in cumulative risk of clinical outcomes using the log-rank test and calculated hazard ratios (HR) and confidence intervals (CI) with the Cox proportional hazards model. We calculated the c-statistic to evaluate the discriminative ability of the clustering algorithm.

*Plasma protein measurements in validation cohort*

In the HFPGR validation cohort, blood samples were collected at enrolment, immediately aliquoted and stored at -70°C for a median of 5.7 years (Q1 4.4; Q3 6.6). Proteomic analyses of EDTA plasma samples were performed using the SomaScan® V4 Assay, a platform for quantifying 5,284 human proteins. The quality control was described previously[2,3], but in brief, systematic biases in raw assay data were corrected following SomaLogic data standardization protocols, involving multiple normalization and calibration steps. These included Hybridization Control Normalization, Intraplate Median Signal Normalization, and Median Signal Normalization to a global reference. Global reference standards were set for serum and plasma matrices, with controls, QC samples, and calibrators on each plate adjusted to these references. Any deviations in assay performance were monitored over time. An overall protein measurement quality metric was calculated for each sample, with all passing the recommended thresholds. The proteomics data was exported as SomaLogic ADAT files, which were imported into R using the *readat* package to remove these proteins with low quality.

### Supplemental results

*Clustering on clinical variables*

The genetic algorithm flagged six clinical characteristics (age, aetiology, and history of coronary artery disease [CAD], arrhythmia, hypertension, and smoking) as relevant for our multivariate clustering, which resulted in three clusters (**Appendix Figure S2AB**). Cluster 1 consisted for 85% of male patients, 94% of patients was diagnosed with CAD, 80% had suffered a myocardial infarction, 42% used antiplatelets, and 87% statins. The patients in cluster 2 were oldest with a median age of 76 years, 88% of the patients was diagnosed with hypertension, 56% with atrial fibrillation, and 66% with renal failure. Cluster 3 consisted of the youngest patients with a median age of 57 years, 69% of the patients had an underlying cardiomyopathy, and patients in this cluster had the lowest burden of comorbidities (**Appendix Table S4**). Based on these differences, the three clusters were mapped to the following clinically relevant HF subclasses: ischemic aetiology (cluster 1), hypertensive aetiology (cluster 2), or underlying cardiomyopathy (cluster 3).

*Combined clustering and sensitivity analysis*

Combined clustering, using the six clinical clustering variables and the first 20 PCs, resulted in three clusters (**Appendix Figure S2EF**). Cluster 1 was almost identical to proteomic cluster 1, with a lower New York Heart Association class (83% I/II) and a low comorbidity burden. Combined clusters 2 and 3 were very different from proteomic clusters 2 and 3. Combined cluster 2 consisted of the oldest patients with a median age of 72 years and the highest comorbidity burden (**Appendix Table S6**).

The sensitivity analysis using LPA resulted in three clusters, with the first cluster consisting of younger individuals with a median age of 64 years (Q1 55; Q3 71) and the highest percentage of females (32%). The second cluster grouped 17 patients, with a median age of

72 years (Q1 68; Q3 74), 12 patients suffering from renal failure (71%) and six from diabetes (35%). The 125 patients in the third cluster had a median age of 65 years (Q1 53; Q3 74) and had the highest New York Heart Association class (37% III/IV). Strikingly, the LVEF was equal among the three clusters (**Appendix Table S13**).

*Cluster membership and outcomes*

Clinical cluster membership did not associate with the clinical outcomes and had a maximum c-statistic of 0.55 (95% CI 0.45; 0.64) for all-cause mortality. Compared to cluster 1, patients in combined cluster 2 and 3 had a very similar increased event rate for all outcomes except for all-cause mortality. Combined cluster 2 was also associated with an increased risk of all-cause mortality (HR 3.15 95% CI 1.19; 8.32). A consistently moderate c-statistic of approximately 0.61 was observed for all outcomes (**Appendix Table S7**).

Taking cluster 1 identified by the LPA analysis as a reference, patients in cluster 3 had an increased event rate for all outcomes, with limited difference comparing outcomes of cluster 2 to cluster 1. The c-statistic ranged between 0.58 and 0.63 for all outcomes (**Appendix Table S14**).

**Supplemental figure legends**

**Figure S1. Cumulative variance explained by the principal components of the proteomic clustering.**

**Figure S2. Bayesian Information Criterion (BIC) for A) clinical, C) proteomic, and E) combined clustering. Log likelihood ratio distribution (BLRT) for B) clinical, D) proteomic, and F) combined clustering.**

NB. Vertical lines represent the observed log likelihood difference with which the p-values were determined.

**Figure S3. Event-free survival per clinical outcome for the A) clinical and B) combined clusters.**

N.B. p-values were calculated using the log-rank test.

**Figure S4. Proportion of clinical characteristics per cluster.**

Abbreviations: AF = atrial fibrillation, CAD = coronary artery disease, MI = myocardial infarction, NYHA = New York Heart Association.

**Figure S5. Association of differentially expressed proteins and canonical cardiac proteins with clinical outcomes.**

The model was derived in 379 participants of the BioSHiFT cohort. Bars represent the 95% confidence interval.

NB. Follow-up was truncated at three years. Abbreviations: CI = confidence interval, HF = heart failure, HR = hazard ratio.

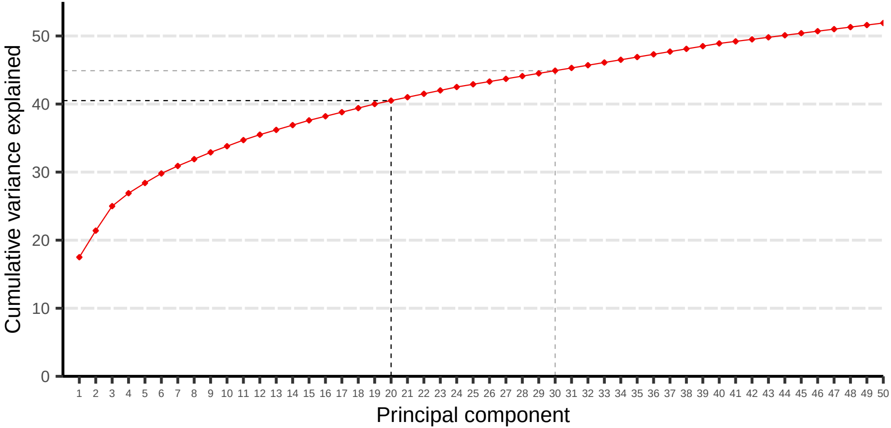

Cumulative variance explained
0
10
20
30
40
50
1 2 3 4 5 6 7 8 9 10 11 12 13 14 15 16 17 18 19 20 21 22 23 24 25 26 27 28 29 30 31 32 33 34 35 36 37 38 39 40 41 42 43 44 45 46 47 48 49 50
Principal component

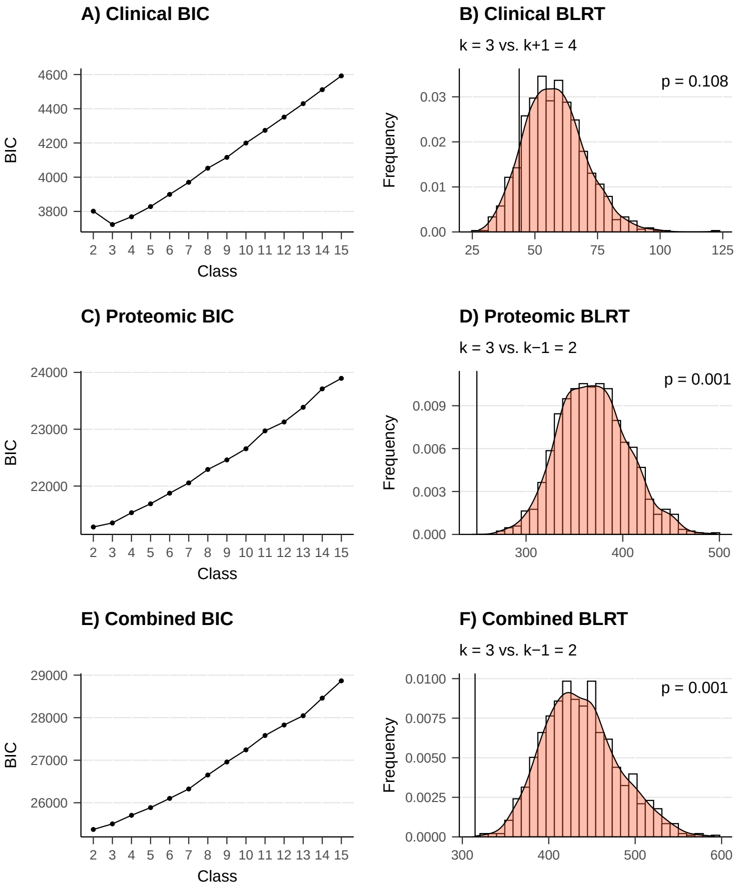

A) Clinical BIC
BIC
Class
B) Clinical BLRT
k = 3 vs. k+1 = 4
p = 0.108
Frequency
C) Proteomic BIC
BIC
Class
D) Proteomic BLRT
k = 3 vs. k−1 = 2
p = 0.001
Frequency
E) Combined BIC
BIC
Class
F) Combined BLRT
k = 3 vs. k−1 = 2
p = 0.001
Frequency

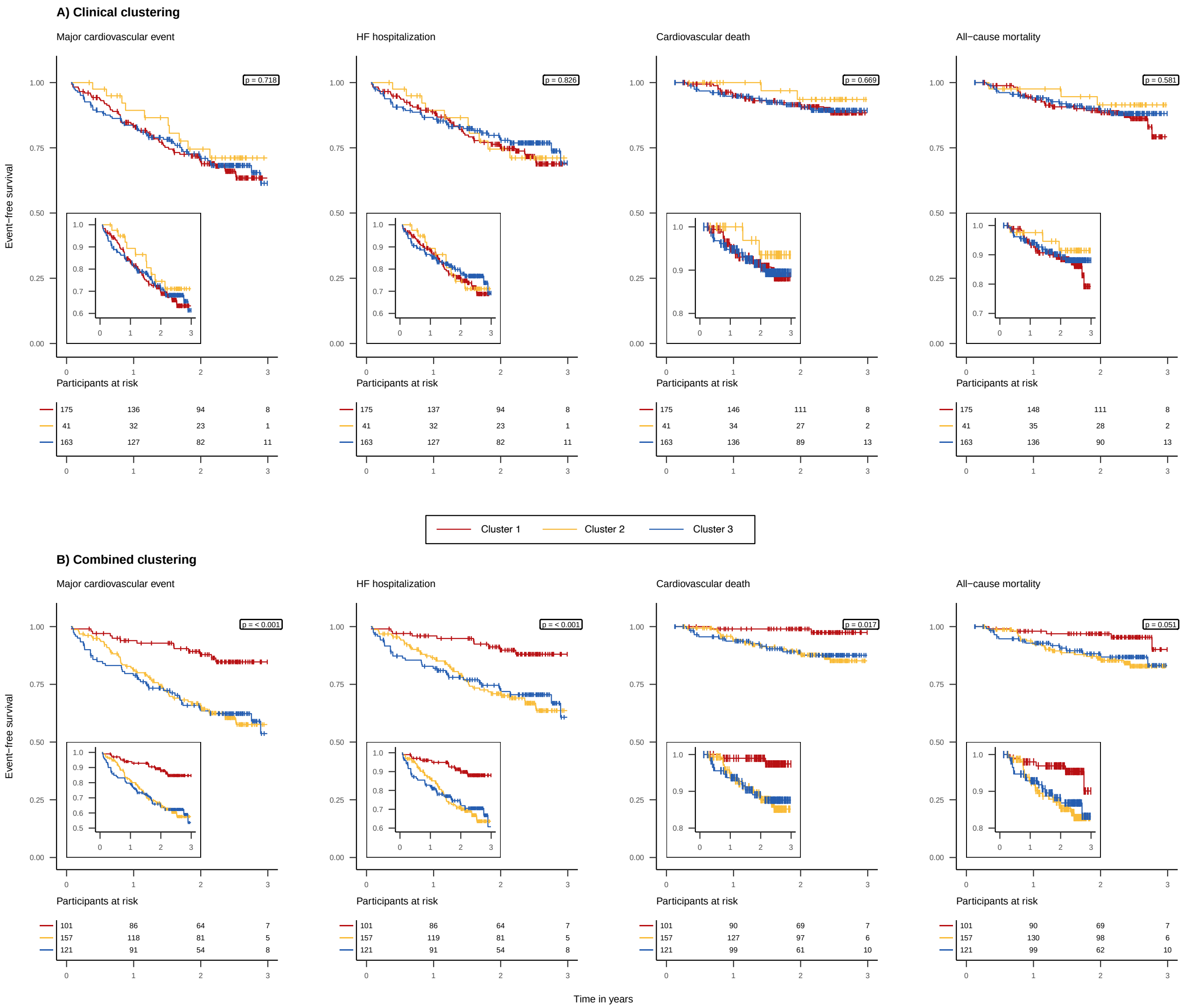
A) Clinical clustering
Major cardiovascular event
p = 0.718
HF hospitalization
p = 0.826
Cardiovascular death
p = 0.669
All−cause mortality
p = 0.581
Event−free survival
Participants at risk
175 136 94 8
41 32 23 1
163 127 82 11
175 137 94 8
41 32 23 1
163 127 82 11
175 146 111 8
41 34 27 2
163 136 89 13
175 148 111 8
41 35 28 2
163 136 90 13
Cluster 1
Cluster 2
Cluster 3
B) Combined clustering
Major cardiovascular event
p = < 0.001
HF hospitalization
p = < 0.001
Cardiovascular death
p = 0.017
All−cause mortality
p = 0.051
Event−free survival
Participants at risk
101 86 64 7
157 118 81 5
121 91 54 8
101 86 64 7
157 119 81 5
121 91 54 8
101 90 69 7
157 127 97 6
121 99 61 10
101 90 69 7
157 130 98 6
121 99 62 10
Time in years

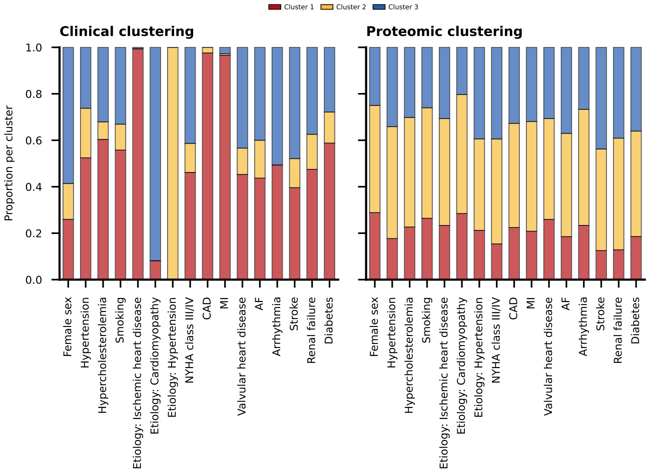

Cluster 1
Cluster 2
Cluster 3
Clinical clustering
Proteomic clustering
Proportion per cluster
1.0
0.8
0.6
0.4
0.2
0.0
Female sex
Hypertension
Hypercholesterolemia
Smoking
Etiology: Ischemic heart disease
Etiology: Cardiomyopathy
Etiology: Hypertension
NYHA class III/IV
CAD
MI
Valvular heart disease
AF
Arrhythmia
Stroke
Renal failure
Diabetes

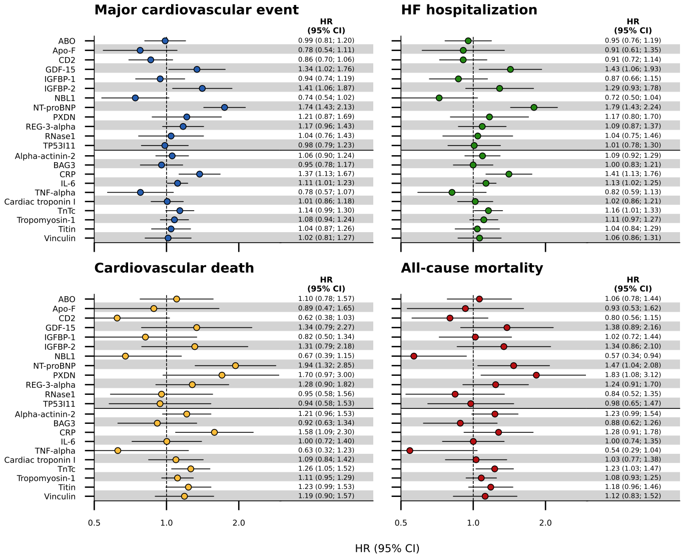

Major cardiovascular event
HR
(95% CI)
ABO 0.99 (0.81; 1.20)
Apo-F 0.78 (0.54; 1.11)
CD2 0.86 (0.70; 1.06)
GDF-15 1.34 (1.02; 1.76)
IGFBP-1 0.94 (0.74; 1.19)
IGFBP-2 1.41 (1.06; 1.87)
NBL1 0.74 (0.54; 1.02)
NT-proBNP 1.74 (1.43; 2.13)
PXDN 1.21 (0.87; 1.69)
REG-3-alpha 1.17 (0.96; 1.43)
RNase1 1.04 (0.76; 1.43)
TP53I11 0.98 (0.79; 1.23)
Alpha-actinin-2 1.06 (0.90; 1.24)
BAG3 0.95 (0.78; 1.17)
CRP 1.37 (1.13; 1.67)
IL-6 1.11 (1.01; 1.23)
TNF-alpha 0.78 (0.57; 1.07)
Cardiac troponin I 1.01 (0.86; 1.18)
TnTc 1.14 (0.99; 1.30)
Tropomyosin-1 1.08 (0.94; 1.24)
Titin 1.04 (0.87; 1.26)
Vinculin 1.02 (0.81; 1.27)
HF hospitalization
HR
(95% CI)
0.95 (0.76; 1.19)
0.91 (0.61; 1.35)
0.91 (0.72; 1.14)
1.43 (1.06; 1.93)
0.87 (0.66; 1.15)
1.29 (0.93; 1.78)
0.72 (0.50; 1.04)
1.79 (1.43; 2.24)
1.17 (0.80; 1.70)
1.09 (0.87; 1.37)
1.04 (0.75; 1.46)
1.01 (0.78; 1.30)
1.09 (0.92; 1.29)
1.00 (0.83; 1.21)
1.41 (1.13; 1.76)
1.13 (1.02; 1.25)
0.82 (0.59; 1.13)
1.02 (0.86; 1.21)
1.16 (1.01; 1.33)
1.11 (0.97; 1.27)
1.04 (0.84; 1.29)
1.06 (0.86; 1.31)
Cardiovascular death
HR
(95% CI)
ABO 1.10 (0.78; 1.57)
Apo-F 0.89 (0.47; 1.65)
CD2 0.62 (0.38; 1.03)
GDF-15 1.34 (0.79; 2.27)
IGFBP-1 0.82 (0.50; 1.34)
IGFBP-2 1.31 (0.79; 2.18)
NBL1 0.67 (0.39; 1.15)
NT-proBNP 1.94 (1.32; 2.85)
PXDN 1.70 (0.97; 3.00)
REG-3-alpha 1.28 (0.90; 1.82)
RNase1 0.95 (0.58; 1.56)
TP53I11 0.94 (0.58; 1.53)
Alpha-actinin-2 1.21 (0.96; 1.53)
BAG3 0.92 (0.63; 1.34)
CRP 1.58 (1.09; 2.30)
IL-6 1.00 (0.72; 1.40)
TNF-alpha 0.63 (0.32; 1.23)
Cardiac troponin I 1.09 (0.84; 1.42)
TnTc 1.26 (1.05; 1.52)
Tropomyosin-1 1.11 (0.95; 1.29)
Titin 1.23 (0.99; 1.53)
Vinculin 1.19 (0.90; 1.57)
0.5
1.0
2.0
All-cause mortality
HR
(95% CI)
1.06 (0.78; 1.44)
0.93 (0.53; 1.62)
0.80 (0.56; 1.15)
1.38 (0.89; 2.16)
1.02 (0.72; 1.44)
1.34 (0.86; 2.10)
0.57 (0.34; 0.94)
1.47 (1.04; 2.08)
1.83 (1.08; 3.12)
1.24 (0.91; 1.70)
0.84 (0.52; 1.35)
0.98 (0.65; 1.47)
1.23 (0.99; 1.54)
0.88 (0.62; 1.26)
1.28 (0.91; 1.78)
1.00 (0.74; 1.35)
0.54 (0.29; 1.04)
1.03 (0.77; 1.38)
1.23 (1.03; 1.47)
1.08 (0.93; 1.25)
1.18 (0.96; 1.46)
1.12 (0.83; 1.52)
0.5
1.0
2.0
HR (95% CI)
